# An improved method to estimate the effective reproduction number of the COVID-19 pandemic: lessons from its application in Greece

**DOI:** 10.1101/2020.09.19.20198028

**Authors:** Theodore Lytras, Vana Sypsa, Demosthenes Panagiotakos, Sotirios Tsiodras

## Abstract

**Introduction:** Monitoring the time-varying effective reproduction number *R*_*t*_ is crucial for assessing the evolution of the COVID-19 pandemic. We present an improved method to estimate *R*_*t*_ and its application to routine surveillance data from Greece.

**Methods:** Our method extends that of Cori et al (2013), adding Bayesian imputation of missing symptom onset dates, imputation of infection times using an external estimate of the incubation period, and an adjustment for reporting delay. To facilitate its use, we provide an R software package named “bayEStim”. We applied the method to COVID-19 surveillance data from Greece, and examined the resulting *R*_*t*_ estimates in relation to control measures applied, in order to assess their effectiveness. We also associated *R*_*t*_, as a measure of transmissibility, to population mobility as recorded in Google data and to ambient temperature. We used a serial interval between 4 and 7.5 days, and a median incubation period of 5.1 days.

**Results:** In Greece *R*_*t*_ fell rapidly as the first control measures were introduced, dropping below 1 at least a week before a full lockdown came into effect. In mid-July *R*_*t*_ started increasing again, as increased mobility associated with tourism activity was observed. Each 10% of increase in relative mobility increased *R*_*t*_ by 8.1% (95% CrI 6.1–10.2%), whereas each unit celsius of temperature increase decreased *R*_*t*_ by 4.6% (95% CrI 5.4–13.7%).

**Conclusions:** Mobility patterns significantly affect *R*_*t*_. Most of the reduction in COVID-19 transmissibility in Greece occurred already before the lockdown, likely as a result of decreased population mobility. Lower viral transmissibility in summer does not appear sufficient to counterbalance the increased mobility due to tourism. Monitoring *R*_*t*_ is an essential component of COVID-19 surveillance, and it is crucial for correctly assessing the effect of control measures.

## Introduction

The coronavirus disease 2019 (COVID-19) pandemic originated in December 2019 in the city of Wuhan, China, spreading across the globe and causing millions of cases and hundreds of thousands of deaths within a few months [1,2]. As the world braces for likely further pandemic waves in late 2020, disease surveillance is crucial in order to monitor the situation and guide public health action. An essential component of COVID-19 surveillance is estimation of the time-varying effective reproduction number *R*_*t*_, defined as the average number of secondary cases at time *t* produced by an infected individual over his/her infectious period. *R*_*t*_ reflects the real-world transmissibility of a pathogen, and is affected by the mobility patterns, social mixing, control measures, population immunity and other factors that are prevalent at each point in time. *R*_*t*_>1 indicates exponential growth, *R*_*t*_=1 indicates sustained transmission and *R*_*t*_<1 suggests exponential decay of an epidemic.

Various methods to estimate *R*_*t*_ have been described in the literature [3–5]; we present an improvement that addresses the inherent limitations of surveillance data, such as incompleteness and reporting delays, allowing for almost real-time estimation of epidemic trends. We apply our method in the case of the COVID-19 pandemic in Greece, and examine how transmissibility of the SARS-CoV-2 virus has varied over time and in relation to control measures, population mobility and ambient temperature.

## Methods

### Estimation of the effective reproduction number R_t_

Our way of estimating *R*_*t*_ extends the methods by Cori et al [3], implemented in a Bayesian framework. In brief, *R*_*t*_ is estimated as the ratio of new locally-acquired infections at time *t*, to the sum of already infected individuals (local or imported) weighted by an infectivity function *w*_*s*_ that is approximated by the serial interval distribution [3,6]. We parametrically model the serial interval with a Gamma distribution, with user-provided mean and standard deviation (SD); a range can be specified for both parameters, from which values are randomly drawn, to include additional uncertainty in the serial interval estimates. As in the original method, in order to reduce noise in the *R*_*t*_ estimates a rolling time window can be specified over which transmission is assumed constant; this is often set at 7 days.

As dates of symptom onset are often partially missing in real-world surveillance data, we use the subset of observed durations between symptom onset and case ascertainment, assumed to also follow a Gamma distribution, to perform Bayesian imputation of any unknown symptom onset dates. This includes asymptomatic cases, for whom we impute an onset date assuming similar duration between infection and case detection as symptomatic cases. In similar fashion, we use a Gamma distribution for the incubation time (again with user-provided mean and SD) to impute infection times for all cases. Whereas using symptom onset dates results in exact but time-lagging estimates of *R*_*t*_ [3], using infection times produces time-accurate estimates, which are necessary in order to assess the effectiveness of control measures and other factors that potentially affect transmission.

Bayesian imputation of infection times incorporates the appropriate uncertainty in the underlying epidemic curve of infections, reflecting the full range of curves that are compatible with the observed data. At the same time this smoothens out both the epidemic curve and the resulting *R*_*t*_ estimates, making it harder to detect abrupt changes in transmissibility and their possible relation to control measures [7]. Although a degree of smoothing is desirable, if this is deemed excessive, a shorter rolling time window can be used.

Delays in case diagnosis and reporting (case ascertainment) will result in right-truncation, i.e. artificially low infection counts for later days in the time series, biasing recent *R*_*t*_ estimates downwards. Several methods have been proposed to adjust for this, which are often referred to as “nowcasting” [8,9]. Our approach was to divide the later counts by the cumulative probability of ascertainment, as given by the distribution of the duration between infection and ascertainment. This simple method has the advantage of not requiring additional historical or other data, compared to more elaborate methods. As an alternative we used a “data augmentation” approach, whereby future cases were added in the dataset based on the reporting rate of the previous week. A comparison of the two approaches is included in the online supplement (Supplementary Figure 1).

**Figure 1:**
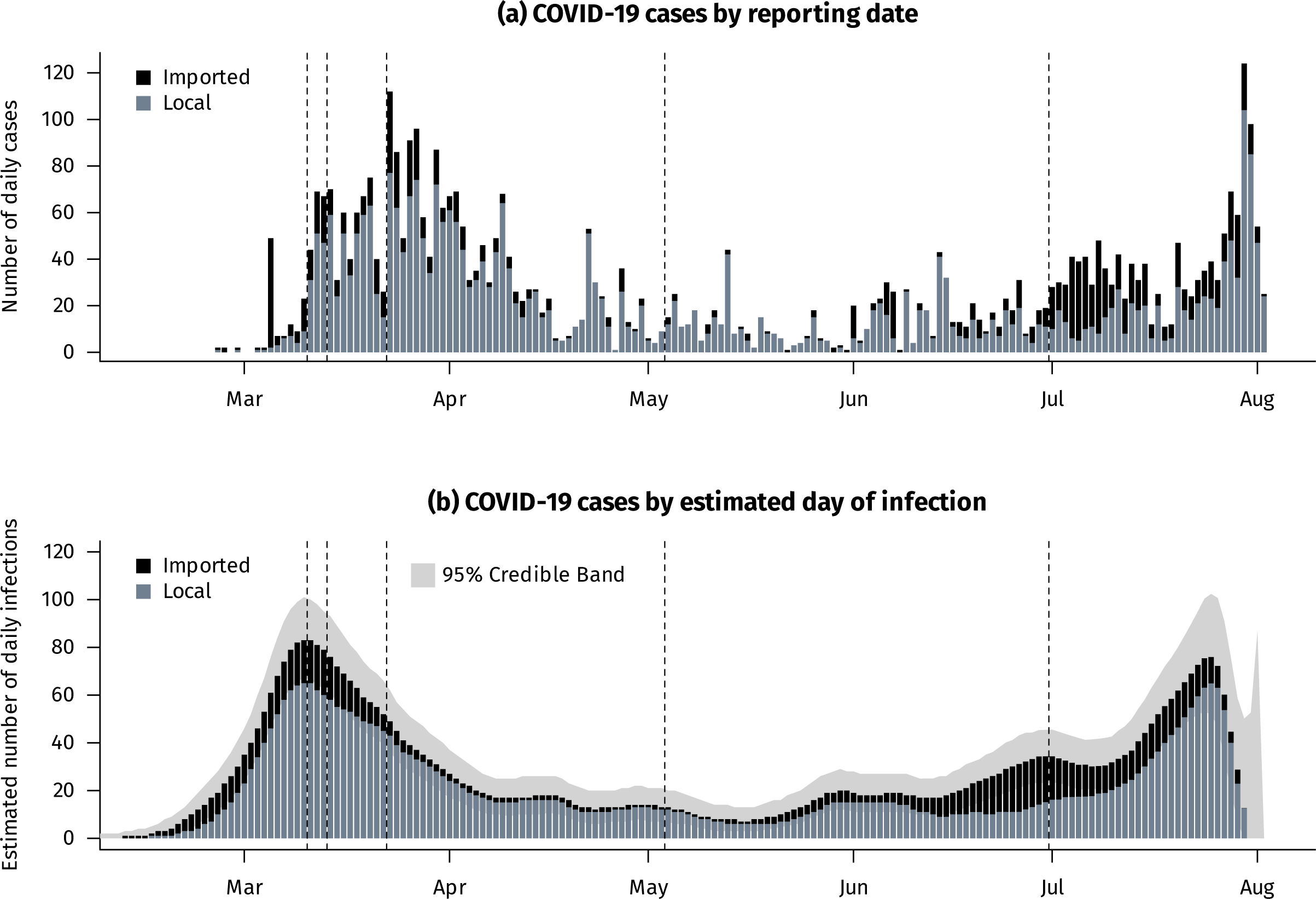
Daily confirmed COVID-19 cases by reporting date and estimated day of infection, Greece, February–August 2020.

The overall model is run using Markov Chain Monte Carlo (MCMC) in JAGS [10]. To facilitate its use, we have created an R software package named “bayEStim” (https://github.com/thlytras/bayEStim) providing a user-friendly interface, in similar fashion to the “EpiEstim” package for the original method [3]. At a minimum the package requires only a vector of symptom onset dates, an indicator of whether the case is local or imported, and a mean and SD for the serial interval. In addition, the dates of ascertainment and the mean and SD of the incubation period can be specified for optimal inference.

### Data sources and analysis

We applied our method for estimating *R*_*t*_ on national-level COVID-19 surveillance data collected in Greece by the National Public Health Organization (NPHO) from 26 February (when the first case was identified) to 3 August 2020. All cases were laboratory-confirmed with RT-PCR testing, and were considered imported if they had arrived in Greece in the last 3 days before a swab was obtained. For the serial interval we specified ranges of 4.0-7.5 days for the mean and 2.0-5.0 days for the SD, in order to cover the estimates reported in the literature [11–13]. Similarly, the mean incubation period was specified as 5.1 days with an SD of 3 days [14]. A 7-day rolling time window was used for *R*_*t*_ estimation. Sensitivity analyses were undertaken with shorter time windows, as well as with the subsets of hospitalized cases and severe cases (defined as hospitalized in intensive care or dead). Dates when major control measures were implemented (or relaxed) were overlaid on the resulting time series of *R*_*t*_ estimates, in order to assess their effects on COVID-19 transmissibility.

In addition, we sought to examine the association between population mobility, ambient temperature and *R*_*t*_ during the study period. We downloaded the Google mobility data for Greece for the study period [15], and averaged the “retail and recreation”, “parks”, “transit stations” and “workplaces” categories into a single relative mobility measure (compared to the baseline period of 3 January to 6 February 2020). As a sensitivity analysis we omitted “parks” from the mobility measure, as it comprises mostly outdoors activities. From the National Oceanic and Atmospheric Administration (NOAA) website [16], we downloaded mean daily temperatures at 48 weather stations across Greece and used a population-weighted average as the overall countrywide temperature of each day. Then, given the 7-day rolling window, we fitted a linear regression of the log*R*_*t*_ estimates on the 7-day moving averages of mobility and temperature. To incorporate the uncertainty of the dependent variable (*R*_*t*_), we ran the regressions separately on each *R*_*t*_ series obtained in each MCMC iteration and combined the regression coefficients using Rubin’s rules [17]. Alternative specifications of this regression were explored as sensitivity analyses. All analyses were undertaken in the R software environment, version 4.0.2 [18].

## Results

Until 3 August 2020 a total of 4,737 COVID-19 cases had been reported in Greece. Of those, 280 cases were from two large clusters neither linked to nor representative of the general population (a large cruiseferry arriving in Greece from abroad, and a migrant hosting facility); these were excluded, leaving 4,459 cases for the *R*_*t*_ estimation. Table 1 presents the age distribution of these cases and their breakdown by location of transmission and severity; approximately one quarter were imported, one third were hospitalized, and 8% were severe cases, i.e. were hospitalized in intensive care or died. Imported cases were on average younger than local cases, while hospitalized and severe cases were older than non-hospitalized (p<0.001, Table 1). A date of symptom onset was reported for 2,510 cases (56.3%).

**Table 1:** Age distribution of laboratory-confirmed COVID-19 cases by location of transmission and severity, Greece, 26 February to 3 August 2020.

|  | **All cases** | **Local** | **Imported** | **Hospitalized** | **Severe** |
| --- | --- | --- | --- | --- | --- |
| Median age (IQR) | 46 (31–60) | 48 (32–62) | 41 (28–53) | 60 (47–73) | 70 (59–81) |
| 0-17 years | 293 (6.6%) | 251 (7.7%) | 42 (3.6%) | 37 (2.6%) | 1 (0.3%) |
| 18-39 years | 1385 (31.1%) | 903 (27.5%) | 482 (41.0%) | 192 (13.7%) | 6 (1.7%) |
| 40-64 years | 1822 (40.9%) | 1367 (41.7%) | 455 (38.7%) | 606 (43.1%) | 123 (34.8%) |
| 65+ years | 819 (18.4%) | 701 (21.4%) | 118 (10.0%) | 569 (40.5%) | 223 (63.2%) |
| Age unknown | 138 (3.1%) | 58 (1.8%) | 80 (6.8%) | 2 (0.1%) | 0 (0.0%) |
| **Total** | 4457 (100.0%) | 3280 (73.6%) | 1177 (26.4%) | 1406 (31.5%) | 353 (7.9%) |

The epidemic curves based on reporting date and estimated date of infection (according to the Bayesian imputation performed in our model) are shown on Figure 1. The lag between infection and reporting is apparent, as is the smoothing introduced by the Bayesian imputation that diminishes the noise of day-to-day reporting. The vertical dashed lines represent the dates when the most important major control measures were introduced or relaxed (see also Supplementary Table 1): on 11 March 2020 schools were closed, on 14 March the food service industry and shopping centres were closed, and on 23 March all non-essential movement was restricted (i.e. a lockdown was implemented). On 4 March the lockdown was lifted, with most business restrictions gradually relaxed until the end of May. And on 1 July international airport connections were restored with most countries, effectively allowing the Greek tourism sector to operate. Although the first peak in daily cases occurred at the time of the lockdown, infections appear to peak much earlier, specifically at the time of school closure. Similarly, a second peak on late August corresponds to an increase of infections since early July, coincidental with an influx of imported cases (Figure 1).

Figure 2 illustrates the *R*_*t*_ estimated by our model, overlaid on the relative mobility and mean daily temperature time series. From a high of 1.50 (95% CrI 1.14–2.08) just before major control measures were introduced, *R*_*t*_ fell rapidly below 1 already one week before the lockdown, reaching 0.68 (95% CrI 0.58– 0.79) on 23 March and staying in this region until May. This rapid decline in *R*_*t*_ mirrors the sharp decline in mobility (more than 50%) recorded in the Google data, which was also sustained until May. A small bump in late May was followed by an *R*_*t*_ consistently below 1 until early July, when the border opening and influx of imported cases was subsequently followed by a rise in local COVID-19 cases and *R*_*t*_ (Figures 1 and 2).

**Figure 2:**
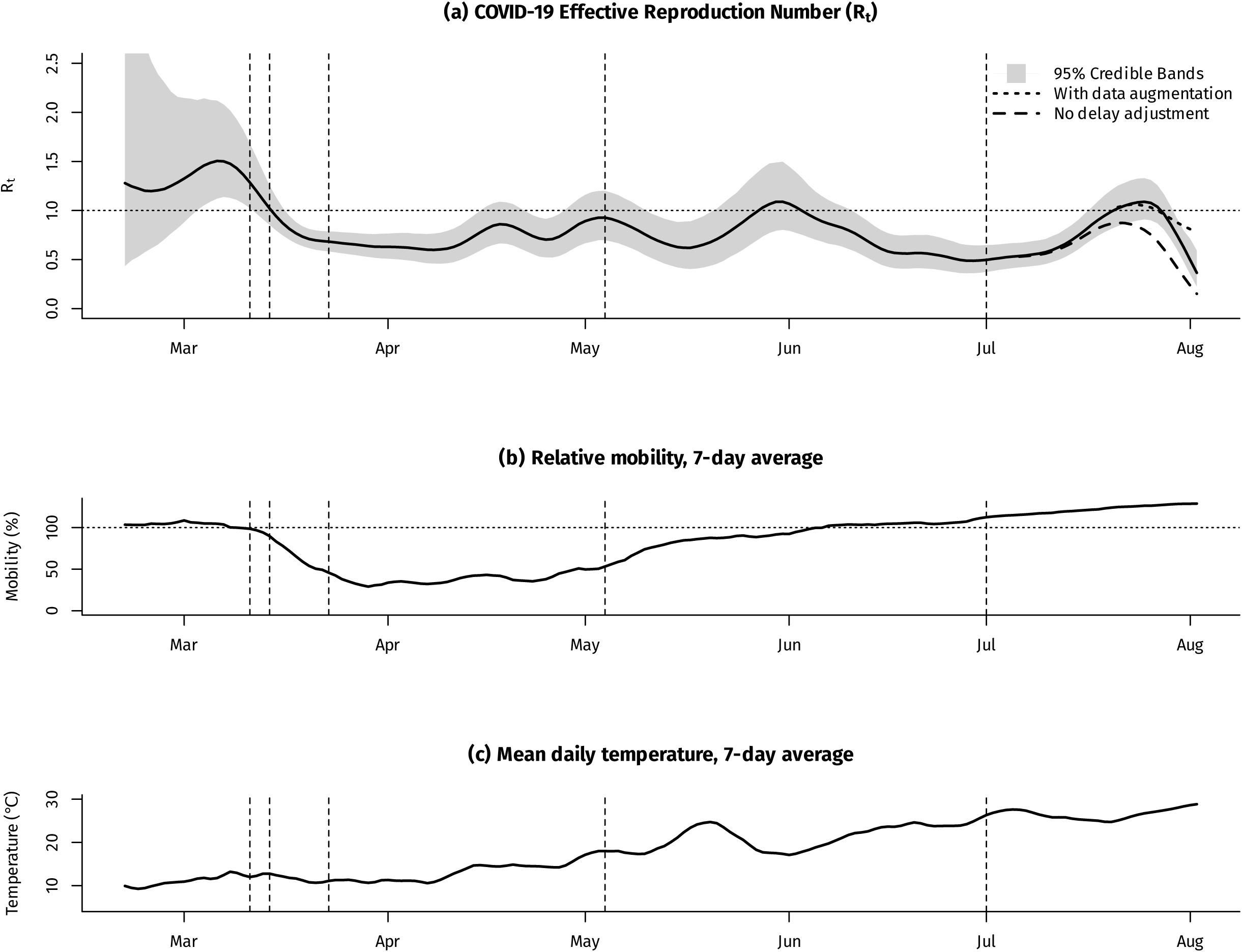
Daily COVID-19 effective reproduction number (*R*_*t*_), relative mobility and mean temperature, Greece, February–August 2020. Footnote for Figure 2: Vertical lines represent the dates when the most important major control measures were introduced or relaxed (see main text for details)

Our delay adjustment method performed relatively well, but consistently underestimated the *R*_*t*_ for the latest 7-10 days in the time series (Figure 2 and Supplementary Figure 1); we therefore omitted the last week of estimates from our regression model (after 27 July). A significant association was found between *R*_*t*_ and both mobility and temperature; each 10% of increase in relative mobility increased *R*_*t*_ by 8.1% (95% CrI 6.1– 10.2%), whereas each unit celsius of increase in the 7-day temperature average decreased *R*_*t*_ by 4.6% (95% CrI 5.4–13.7%). The adjusted R-squared of the model was 54.3%, indicating a fairly good fit to the data, and had the lowest AIC of all alternative models explored as sensitivity analyses (Supplementary Table 2).

In sensitivity analyses using hospitalized and severe cases, the *R*_*t*_ time series was very similar to the one obtained using all cases, albeit with much reduced precision given the lower numbers of cases (Supplementary Figure 2).

## Discussion

Monitoring the transmissibility of the SARS-CoV-2 virus during the COVID-19 pandemic is crucial in order to assess the effectiveness of control measures and guide public health policy. *R*_*t*_ is a useful and easily understandable metric for this purpose, but its reliable estimation presents considerable challenges [7], especially given the inherent limitations of surveillance data [19]. Our method to estimate *R*_*t*_ does not require structural assumptions other than the serial interval distribution, and introduces several improvements that make it appropriate for use in a surveillance context. It works with minimal or incomplete data, it largely adjusts for reporting delays, and can produce time-accurate estimates using an external estimate of the incubation period; the latter is essential for correctly assessing the effectiveness of control measures, as any intervention to reduce infection rates will only be reflected in case reporting rates after a substantial period of time. Our method is also very easy to use through our “bayEStim” package for the R software environment.

On the other hand, there are certain limitations with our approach. Imprecision can be high especially with low case counts, as our model incorporates all sources of uncertainty in *R*_*t*_, serial interval, incubation period, missing symptom onset dates and delay distribution. Bayesian imputation of infection times also introduces smoothing, which is both appropriate and to an extent desirable, but can also blur abrupt changes in *R*_*t*_ [7]. This should be kept in mind when interpreting the results. Substantial changes in testing or ascertainment rates over time can bias the results, as can the presence of few large clusters compared to overall cases; down-weighting of cluster-related cases, for example according to their percentage positive compared to overall cases, may be a possible solution and has been implemented in “bayEStim”. Ideally our method should be applied to hospitalized or severe case series, which can be less subject to underascertainment or non-random testing, although in our case *R*_*t*_ estimates were not appreciably different.

Applying our method of *R*_*t*_ estimation to Greek COVID-19 surveillance data yielded several important results, with clear implications for the past and future management of the pandemic. First, there was a clear relationship between population mobility and viral transmissibility, indicating that social distancing and staying home can be very effective in bringing the *R*_*t*_ of COVID-19 below 1 [20]. Indeed both mobility and Rt started falling rapidly just as the first control measures were introduced, indicating that the multiple control measures introduced, combined with good public compliance, resulted in effective social distancing earlier than the full lockdown of 23 March. This was likely crucial, as the virus did not get enough time to spread widely among the population. Counterintuitively, the lockdown appears to not have substantially contributed to a further reduction in COVID-19 transmissibility, as *R*_*t*_ had already fallen significantly below 1 at least a week earlier and remained stable thereafter. It is possible that without the lockdown the decrease in mobility and *R*_*t*_ might not have been sustained, but this purely hypothetical.

As social distancing measures were relaxed, *R*_*t*_ in Greece remained largely below 1 until early July. Then a large influx of tourists (with 1.3 million arrivals during all of July [21], in a population of around 10.5 million), with hundreds of imported COVID-19 cases (Figure 1), was followed by a steep increase in locally-acquired cases and *R*_*t*_ climbing back to 1. This coincided with a continued increase in mobility beyond baseline levels (Figure 2). It should be emphasized that the arrival of imported cases from abroad does *not* in itself raise *R*_*t*_, as transmissibility depends entirely on local conditions of social mixing and mobility. However, summer in Greece is defined by people, Greek and foreign, going on vacation in large numbers and in proximity to each other, especially in tourist hotspots; moreover, an estimated 850,000 local jobs are directly or indirectly linked to the tourism industry [22]. This situation creates the general conditions for sustained spread of the virus among the population, through increased mobility and crowding. It is the combination of increased case importations and increased transmissibility that results in more COVID-19 cases identified during the summer, a phenomenon which is expected to abate in September as the tourist season winds down. In the meantime, is is important to push *R*_*t*_ back below 1 as much as possible, with targeted social distancing measures and widespread use of other protective measures like masks and hand hygiene.

Finally, we found a relationship between ambient temperature and COVID-19 transmissibility; in fact the 18.3 degrees celsius difference between the lowest and highest temperature in our data corresponds to a 58.1% change in *R*_*t*_ (95% CrI 35.7–72.7%), which appears quite significant. This may be attributed to the effects of heat on the virus itself, but is likely more down to behavioural patterns in the population, with more social interaction happening outdoors as the weather gets warmer and vice versa. This effect however does not appear sufficient to counterbalance the increased mobility associated with the summer season in Greece, which is unsurprising considering the lack of population immunity to SARS-CoV-2 [23]. Moreover, with the higher transmissibility anticipated during the winter, a substantially higher degree of social distancing will likely be required to keep *R*_*t*_ below 1 and the pandemic under control.

In conclusion, we present an improved method and software for estimating *R*_*t*_ that is tailored to the realities of a disease surveillance context, and can be a valuable component of the surveillance activities during the current COVID-19 pandemic. Applying our method to the Greek case demonstrates the important conclusions that can be drawn, and that can guide the further management of the pandemic.

## Data Availability

Data are available upon reasonable request to the corresponding author

## Bibliography

1. Wu F, Zhao S, Yu B, Chen Y-M, Wang W, Song Z-G, et al. A new coronavirus associated with human respiratory disease in China. Nature. 2020;579(7798):265–9.

2. Rapid Risk Assessment: Coronavirus disease 2019 (COVID-19) in the EU/EEA and the UK – eleventh update: resurgence of cases [Internet]. European Centre for Disease Prevention and Control. 2020 [cited 2020 Aug 12]. Available from: https://www.ecdc.europa.eu/en/publications-data/rapid-risk-assessment-coronavirus-disease-2019-covid-19-eueea-and-uk-eleventh

3. Cori A, Ferguson NM, Fraser C, Cauchemez S. A new framework and software to estimate time-varying reproduction numbers during epidemics. Am J Epidemiol. 2013 Nov 1;178(9):1505–12.

4. Wallinga J, Lipsitch M. How generation intervals shape the relationship between growth rates and reproductive numbers. Proc Biol Sci. 2007 Feb 22;274(1609):599–604.

5. Bettencourt LMA, Ribeiro RM. Real time bayesian estimation of the epidemic potential of emerging infectious diseases. PLoS ONE. 2008 May 14;3(5):e2185.

6. Thompson RN, Stockwin JE, van Gaalen RD, Polonsky JA, Kamvar ZN, Demarsh PA, et al. Improved inference of time-varying reproduction numbers during infectious disease outbreaks. Epidemics. 2019;29:100356.

7. Gostic KM, McGough L, Baskerville E, Abbott S, Joshi K, Tedijanto C, et al. Practical considerations for measuring the effective reproductive number, Rt. medRxiv. 2020 Jun 20;

8. Höhle M,an der Heiden M. Bayesian nowcasting during the STEC O104:H4 outbreak in Germany, 2011. Biometrics. 2014 Dec;70(4):993–1002.

9. McGough SF, Johansson MA, Lipsitch M, Menzies NA. Nowcasting by Bayesian Smoothing: A flexible, generalizable model for real-time epidemic tracking. PLoS Comput Biol. 2020;16(4):e1007735.

10. Plummer M. JAGS: A program for analysis of Bayesian graphical models using Gibbs sampling. In Vienna; 2003. p. 125.

11. Bi Q, Wu Y, Mei S, Ye C, Zou X, Zhang Z, et al. Epidemiology and transmission of COVID-19 in 391 cases and 1286 of their close contacts in Shenzhen, China: a retrospective cohort study. The Lancet Infectious Diseases [Internet]. 2020 Apr 27 [cited 2020 Apr 30];0(0). Available from: https://www.thelancet.com/journals/laninf/article/PIIS1473-3099(20)30287-5/abstract

12. Du Z, Xu X, Wu Y, Wang L, Cowling BJ, Meyers LA. Serial Interval of COVID-19 among Publicly Reported Confirmed Cases. Emerging Infect Dis. 2020 Mar 19;26(6).

13. Liu T, Qi L, Yao M, Tian K, Lin M, Jiang H, et al. Serial Interval and Reproductive Number of COVID-19 Among 116 Infector-infectee Pairs — Jingzhou City, Hubei Province, China, 2020. CCDCW. 2020 Jul 1;2(27):491–5.

14. Lauer SA, Grantz KH, Bi Q, Jones FK, Zheng Q, Meredith HR, et al. The Incubation Period of Coronavirus Disease 2019 (COVID-19) From Publicly Reported Confirmed Cases: Estimation and Application. Ann Intern Med. 2020 Mar 10;

15. Google. COVID-19 Community Mobility Reports [Internet]. COVID-19 Community Mobility Reports. [cited 2020 Aug 1]. Available from: https://www.google.com/covid19/mobility?hl=en

16. National Oceanic and Atmospheric Administration (NOAA). National Centers for Environmental Information (NCEI) Climate Data Online Search [Internet]. [cited 2020 Aug 1]. Available from: https://www.ncdc.noaa.gov/cdo-web/search

17. Rubin DB. Multiple imputation for nonresponse in surveys. New York: John Wiley & Sons; 1987.

18. R Core Team. R: A Language and Environment for Statistical Computing [Internet]. Vienna, Austria: R Foundation for Statistical Computing; 2019. Available from: http://www.R-project.org/

19. Giesecke J. Routine surveillance of infectious diseases. In: Modern infectious disease epidemiology. CRC Press; 2017.

20. Sypsa V, Roussos S, Paraskevis D, Lytras T, Tsiodras S, Hatzakis A. Modelling the SARS-CoV-2 first epidemic wave in Greece: social contact patterns for impact assessment and an exit strategy from social distancing measures. medRxiv. 2020 May 29;2020.05.27.20114017.

21. Greek City Times. Number Of Imported Coronavirus Cases Remains Low, Says Greek Tourism Minister [Internet]. Greek City Times. [cited 2020 Aug 15]. Available from: https://greekcitytimes.com/2020/08/07/number-of-imported-coronavirus-cases-remains-low-says-greek-tourism-minister/

22. ekathimerini.com. WTTC praises Greece’s 2019 tourism growth and response to Covid-19 [Internet]. [cited 2020 Aug 16]. Available from: http://www.ekathimerini.com/252473/article/ekathimerini/business/wttc-praises-greeces-2019-tourism-growth-and-response-to-covid-19

23. Bogogiannidou Z, Vontas A, Dadouli K, Kyritsi MA, Soteriades S, Nikoulis DJ, et al. Repeated leftover serosurvey of SARS-CoV-2 IgG antibodies, Greece, March and April 2020. Euro Surveill. 2020;25(31).

